# Identification of a Novel *SLC2A9* Gene Association with LDL-C levels and Evaluation of Polygenic Scores in a Multi-Ancestry Genome Wide Association Study

**DOI:** 10.1101/2024.07.04.24309936

**Authors:** Umm-Kulthum Ismail Umlai, Salman M. Toor, Yasser A. Al-Sarraj, Shaban Mohammed, Moza S H Al Hail, Ehsan Ullah, Khalid Kunji, Ayman El-Menyar, Mohammed Gomaa, Amin Jayyousi, Mohamad Saad, Nadeem Qureshi, Jassim M Al Suwaidi, Omar M E Albagha

**Affiliations:** College of Health and Life Sciences (CHLS), Hamad Bin Khalifa University (HBKU), Qatar Foundation (QF), P.O. Box 34110, Doha, Qatar; Qatar Genome Program (QGP), Qatar Foundation Research, Development and Innovation, Qatar Foundation, P.O. Box 5825, Doha, Qatar; Department of Pharmacy, Hamad Medical Corporation, P.O. Box 3050, Doha, Qatar; Qatar Computing Research Institute (QCRI), Hamad Bin Khalifa University (HBKU), Qatar Foundation (QF), P.O. Box 34110, Doha, Qatar; Trauma and Vascular Surgery, Hamad Medical Corporation (HMC), P.O. Box 3050, Doha, Qatar; Adult Cardiology, Heart Hospital, Hamad Medical Corporation (HMC), P.O. Box 3050, Doha, Qatar; Department of Diabetes, Hamad Medical Corporation (HMC), P.O. Box 3050, Doha, Qatar; Primary Care Stratified Medicine Research Group, Centre for Academic Primary Care, School of Medicine, University of Nottingham, Nottingham, NG7 2UH, United Kingdom; Centre for Genomic and Experimental Medicine, Institute of Genetics and Cancer, University of Edinburgh, Edinburgh, EH4 2XU, United Kingdom

## Abstract

The genetic determinants of low-density lipoprotein cholesterol (LDL-C) levels in blood have been predominantly explored in European populations and remain poorly understood in Middle Eastern populations. We investigated the genetic architecture of LDL-C variation in the Middle Eastern population of Qatar. Whole genome sequencing data of 13,701 individuals (discovery; n=5939, replication; n=7762) from the population-based Qatar Biobank (QBB) cohort was used to conduct a genome-wide association study (GWAS) on serum LDL-C levels. We replicated 168 previously reported loci from the largest LDL-C GWAS conducted by the Global Lipids Genetics Consortium (GLGC), with high correlation in allele frequencies (R^2^=0.77) and moderate correlation in effect sizes (Beta; R^2^=0.53). We also performed a multi-ancestry meta-analysis with the GLGC study using MR-MEGA (Meta-Regression of Multi-Ethnic Genetic Association). The multi-ancestry meta-analysis identified one novel LDL-C-associated locus; rs10939663 (*SLC2A9*; genomic control-corrected *P*=1.25×10^−8^). Lastly, we developed Qatari-specific polygenic score (PGS) panels from our discovery dataset and tested their performance in the replication dataset against PGS derived from other ancestries. The multi-ancestry derived PGS (PGS000889) performed best at predicting LDL-C levels, whilst the Qatari-derived PGS panels also showed comparable performance. Overall, we report one novel gene variant associated with LDL-C levels, which may be explored further to decipher its potential role in the etiopathogenesis of cardiovascular diseases. Our findings also highlight the importance of population-based genetics in developing PGS panels for clinical utilization.

## Introduction

Cardiovascular diseases (CVD) remain the leading cause of global mortality. Elevated low-density lipoprotein cholesterol (LDL-C) concentration in blood, which is a known heritable and modifiable risk factor for the development of atherosclerotic CVD, presents as an increasingly prevalent global health burden (1, 2). Notably, the incidence of CVD is on the rise in the Middle Eastern region (3). The incidence of death per 100,000 individuals attributed to CVD is also higher in the Middle East compared to Europe and USA, and predominantly affects males aged 58–59 years, which is a younger age band compared to the rest of the world (4).

An elevated level of low-density lipoprotein cholesterol (LDL-C) is identified as one of the main risk factors for the development of CVD, and lowering LDL-C concentration leads to a significant reduction in major vascular and coronary complications and stroke (5, 6). Deciphering the genetic determinants of variation in LDL-C levels has merits of improving the understanding of LDL-C regulation. Additionally, it has the potential for identifying novel molecular targets and for resolving the paradigm around the development and progression of atherosclerotic CVD. Pathogenic variants in genes involved in LDL-C uptake and catabolism such as *LDLR*, *APOB*, *APOE*, *LDLRAP1* and *PCSK9* are recognized as the principal causative factors for monogenic Familial Hypercholesterolemia (FH), one of the most common inherited autosomal condition in the world (7). A recent study showed that the prevalence of FH was estimated at 1 in 125 individuals in the Middle eastern population of Qatar which is higher than the global prevalence (26). However, variation in LDL-C concentration is a quantitative trait that is broadly dictated by a polygenic cause due to common alleles with small/moderate effect, which influence multiple LDL-C modulating genes that can lead to raised LDL-C levels.

Genome-wide association studies (GWAS) have enabled the identification of several genetic risk loci that are associated with blood lipid concentrations (8). The GWAS catalog currently lists over 1,850 genomic risk loci associated with blood lipids (9). Notably, a recent comprehensive GWAS conducted by the Global Lipids Genetics Consortium (GLGC), which represents the largest published GWAS and meta-analysis on LDL-C levels (Graham *et. al.,*) was performed on a cohort of around 1.65 million individuals (10). The study is an aggregate of 201 studies from 5 ancestries: African (6.0%), East Asian (8.9%), European (79.8%), South Asian (2.5%) and Hispanic (2.9%), with little representation of Middle Eastern populations. This large-scale multi-ancestry meta-analysis revealed more than 900 lipid-related genetic loci (10).

The inclusion of additional underrepresented ancestries, such as those of Arab descent, can improve the understanding of lipid level genetic determinants, reveal novel drug targets, advance fine mapping of variants associated with lipid levels and progress polygenic disease prediction (10). Associations of genetic variants with lipid traits and increased susceptibility to CVD are recurrently explored for the development of polygenic scores (PGS) (11–13). The earliest LDL-C PGS developed is a 12-SNP scoring panel from the GLGC, in 2013 (14). This scoring panel was later refined in another study by the same group, developing a 6-SNP score which performed equally well at discriminating between hypercholesterolemia patients with no confirmed mutation and healthy controls (15). This score has not been published on the Polygenic Score Catalog (PGS catalog) (16) but the PGS panel is available in the publication (15).

However, since GWAS have been predominantly conducted on individuals of European ancestry (17), the portability of genetic risk loci among different ancestries requires thorough investigations. This is due to differences in genetic architectures encompassing differences in allele frequencies and linkage disequilibrium (LD) that can affect genetic associations (18). For instance, genetic risk scores for lipid traits developed in European populations did not perform well in Sub-Saharan African populations (19, 20). Population-based GWAS of underrepresented regions can reveal both common and rare genetic variants related to inherited risk of elevated LDL-C concentration and increased susceptibility to CVD, while multi-ancestry meta-analyses may also be advantageous for uncovering novel variants. Increasing diversity in LDL-C GWAS as opposed to mitigating the limitations of currently devised PGS has merits for improving the overall utilization of PGS prediction.

PGS development for predicting LDL-C levels or CVD within the wider Middle Eastern region based on local genomic variation has not been systematically explored. A recent study on a cohort of coronary heart disease (CHD) from Qatar showed that performance of PGS derived from European studies of CVD was comparable to the performance reported in Europeans (21). However, European-derived polygenic score for many quantitative traits performed lower when assessed in the Qatari population compared to European populations. This was revealed in a GWAS conducted in Qatar, which focused on 45 clinically relevant traits (22).

In this current study, we conducted a comprehensive GWAS of LDL-C in the Middle Eastern population of Qatar based on whole genome sequencing (WGS) data of 13,701 individuals from the Qatar Biobank (QBB) cohort. We also performed a multi-ancestry meta-analysis of our GWAS data with the largest published data from GLGC consortium to broaden the spectrum of LDL-C associated loci. Importantly, we derived Qatari-specific PGS panels and assessed their performance to predict LDL-C levels against PGSs derived from African, European and multi-ancestry populations. Overall, our findings contribute towards diversifying the current knowledge of population genetics for LDL-C levels and aid the clinical application of PGS. This study represents the largest GWAS of Arab populations performed on LDL-C association and polygenic risk score panels derived from the largest discovery and replication cohort within Arab populations.

## Methods

### Participant recruitment and data collection

Study participants were recruited from the Qatar Biobank (QBB), a national biorepository, which collects biological samples and clinicopathological data from Qatari citizens in coordination with the national public healthcare provider (Hamad Medical Corporation, Doha, Qatar). Our study cohort comprised of 13,808 subjects aged 18 to 89 years. The first data release of QBB was used as a discovery dataset containing 6,013 participants and the replication dataset was based on the second release comprising of 7,795 subjects. All study participants provided written informed consent prior to inclusion in this study. This study was approved by the institutional review boards of QBB (Protocol no. E-2020-QF-QBB-RES-ACC-0154-0133) and Hamad Bin Khalifa University (HBKU), Doha, Qatar (Approval no. QBRI-IRB 2021-03-078).

Biological samples (blood, urine and saliva) were collected from study participants. In addition, routine medical examination and biochemical tests (e.g. lipid profile, trace mineral concentrations, etc.) were also performed and clinical data retrieved. Participants also filled out medical questionnaires with questions pertaining to medical and family histories, medication, and supplement (23). Cholesterol treatment was determined based on self-reported data from questionnaires.

### LDL-C measurement

LDL-C levels were calculated from blood lipid profiles (Total cholesterol, High-density lipoprotein-cholesterol and Triglycerides) using the Friedewald formula (FF) and recorded in mg/dL. Serum lipid profiles were determined using enzymatic colorimetric assays performed using Roche COBAS 6000 c analyzer (Roche, Switzerland).

### Whole Genome Sequencing

Whole genome sequencing (WGS) was conducted by the Qatar Genome Program (QGP) (24), as previously described (22). Briefly, genomic DNA was isolated from whole blood using the QIAamp DNA Blood MIDI kit (Qiagen, Germany) and QIASymphony SP automated instrument (Qiagen, Hilden, North Rhine-Westphalia, Germany) as per the manufacturer’s instructions. DNA quality was evaluated by the Caliper Labchip GXII (Perkin Elmer, Waltham, MA, USA) Genomic DNA assay and concentrations were measured using the Quant-iT dsDNA Assay (Invitrogen, Waltham, MA, USA). Libraries for WGS were prepared using the Illumina TruSeq DNA Nano kit (Illumina, San Diego, CA, USA). The genomic libraries were sequenced using the HiSeq X Ten (Illumina) at 30X coverage at the Clinical Genomic Facilities at Sidra (Sidra Medicine, Doha, Qatar)

### Quality control and data preprocessing

FastQC (v0.11.2) was used to perform quality control measures on the genomic data files and read alignment was performed against the GRCh38 genome reference. Picard (v1.117; (CollectWgsMetrics) was used to control the quality of mapped reads and the variants were joint called in a combined variant call file (gVCF) for the entire cohort. Downstream quality control and data preprocessing for GWAS analysis preparation were performed using the PLINK tool (v1.9) (25). Genetic variants with call rates < 90%, minor allele frequency (MAF) < 0.01, mean depth coverage <10X, those with a Hardy-Weinberg equilibrium *P*<1.0×10^−6^ were excluded from the analysis. All samples with missing LDL-C level data, excess heterozygosity, mismatched gender, population outliers determined by multi-dimensional scaling were excluded from the dataset. After quality control, the total number of subjects used in downstream analysis was 13,701 comprising 5,939 in the discovery set and 7,762 in the replication set.

### GWAS analysis

LDL-C values were adjusted for age, age^2^, the first ten genetic principal components (PC1 to PC10) and cholesterol treatment separately by gender and residuals were calculated. GWAS was performed using SAIGE (26), applying inverse normalization of the LDL-C residuals and default adjustment for relatedness in the data. To assess replication of known loci, a list of variants from the largest published GWAS of LDL-C by Graham *et. al.,* (10) was downloaded from the University of Michigan Center for Statistic Genetics (http://csg.sph.umich.edu/willer/public/glgc-lipids2021/results/trans_ancestry/). This list was filtered to only include variants meeting the genome-wide significance threshold (*P*<5.0×10^−8^) and then compared with the variants from the GWAS of QBB discovery dataset with nominal significance (*P*<0.05) to identify replicated variants. The R intersect function was used to compare data frames and output common SNP entries. The effect allele frequency and effect size (beta) from the multi-ancestry data of Graham *et. al.,* study was compared to that from our GWAS results using Spearman’s correlation. Effect values and allele frequencies were then plotted using GraphPad PRISM (v9.0; GraphPad software, San Diego, CA, USA).

### Multi-Ancestry Meta analysis

We first performed a fixed effect meta-analysis of QBB discovery and replication GWAS using the METAL (27) tool. Then a multi-ancestry meta-analysis was performed using the results from METAL and with summary statistics from each of the single-ancestry LDL-C GWAS; African (AFR), East Asian (EAS), European (EUR), Hispanic (HIS), South Asian (SAS) from the GLGC study (10). Multi-ancestry Meta-analysis was performed using the Meta-Regression of Multi-AncEstry Genetic Association tool (MR-MEGA (v0.2)) (28) using 3 PCs and genome control correction (GC) correction. Novel variants were identified based on attaining genome-wide significance (GC corrected *P*≤5.0×10^−8^) post-meta-analysis, being beyond 500kb of any previously reported genome-wide significant SNPs from the GLGC study and not in LD with any previously reported genome-wide significant loci within 500kB in the Common Metabolic Disease (CMD) Knowledge Portal (29) or in the GWAS Catalog (9). Beta effect values were calculated by running multi-ancestry meta-analysis using METAL. Regional association plots were generated using the LocusZoom(v1.4) tool (30).

### Analysis of polygenic scores

Polygenic score panels for LDL-C were all derived based on the QBB discovery dataset (training cohort) using three different methods; thresholding-only, clumping and thresholding (C+T) or Bayesian shrinkage implemented in LDpred2 (31). For thresholding, eight different *P*-value thresholds were used, ranging from 5.0×10^−4^ to 5.0×10^−16^. For C+T, GWAS results from the training set were pruned using the --clump option in PLINK (v1.9) at two linkage disequilibrium (LD) clumping r^2^ cutoff values; 0.2 and 0.8, clumping distance of 500 kb and the same *P*-values cut off used in the thresholding method. Only index SNPs from the output of clumping were selected for the scoring panels. The effect size values were obtained from the beta-effect weights from the GWAS of the discovery cohort.

LDpred-2 is a Bayesian shrinkage algorithm from the bigsnpr package on R (32), which was used to derive polygenic scores from the discovery cohort. The LDpred2 ‘grid’ method was used for beta effect size estimation and prediction using the default parameters. A total of 168 different panels were tested by Ldpred-2 and the 3 best performing panels were extracted for assessment in the QBB replication cohort.

PGS scoring for all panels were performed using “--score” and “sum” option in PLINK (v1.9). PGS panels obtained from other studies, derived from European (PGS000892, PGS000893 (10), EUR_6SNP (15), PGS000814 (14), PGS000875 (33), PGS000749 (34)), Multi-ethnic (PGS000888, PGS000889 (10), African (PGS000886, PGS000887) (10) were downloaded from the Polygenic Score catalog (PGS catalog) or associated publication, and used to score the replication cohort.

The performance of PGS panels in predicting LDL-C was assessed using linear regression models with PGS, age, gender, cholesterol treatment, and the first four principal components (PCs) as predictors. Linear regression was performed using R software (v4.3.1) and the resulting adjusted R^2^ values were used as performance metrics.

## Results

### Characteristics of study cohorts

An overview of the overall study design is depicted in Figure 1. After quality control measures, the study cohort comprised 5,939 subjects in the discovery set and 7,762 subjects in the replication set (Total n=13,701). Characteristic features of study cohorts are listed in Supplementary Table 1.

**Figure 1.**
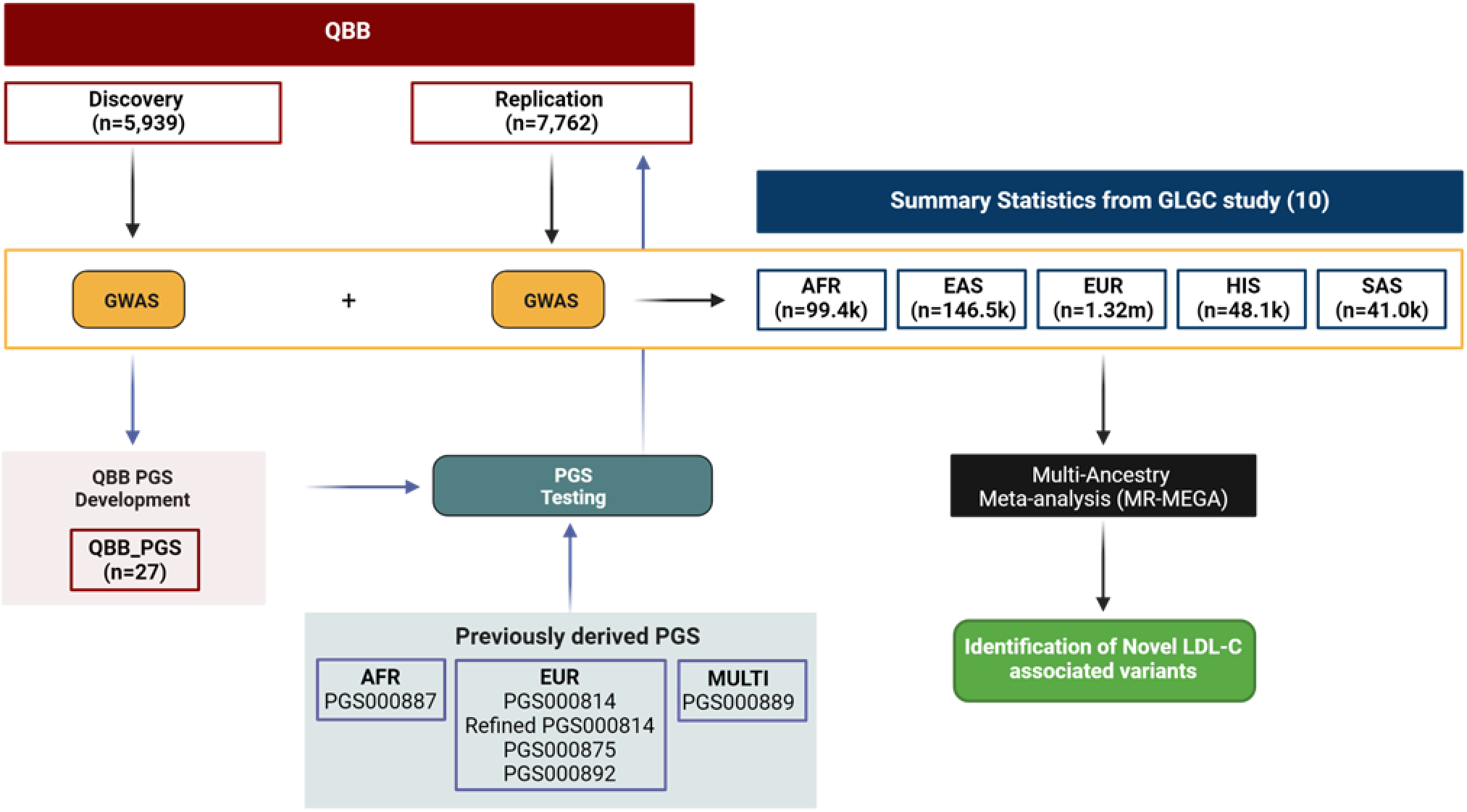
Study design and analysis workflow. LDL-C GWAS was performed on the Qatar Biobank (QBB) discovery (n=5,939) and replication (n=7,762) cohorts separately using inverse-normalized LDL-C residuals. GWAS summary statistics from the QBB cohorts were then combined in a single-ancestry, fixed-effect meta-analysis using METAL. The results were further meta-analyzed using (MR-MEGA-v0.2) with 5 single-ancestry cohort summary statistics data (African; AFR, East-Asian: EAS, European; EUR, Hispanic; HIS and South Asian; SAS) from the GLGC LDL-C study (10). GWAS results from the discovery cohort were then used to develop multiple PGS panels using different tools (refer to methods section). 27 QBB-derived PGS panels (QBB_PGS) were then tested on the QBB replication cohort. Previously derived PGS from other ancestries (AFR (10), EUR (10, 14, 15, 31, 32) and Multi-ancestry; MULTI (10,33)) were also tested in the QBB replication cohort.

### LDL-C GWAS

Manhattan and QQ-plots derived from the GWAS performed for the discovery dataset are shown in Figure 2. The Manhattan plot illustrates 5 genome-wide significant loci located in chromosome bands 1p13.3, 2p24.1, 19q13.32, 19p13.2 and 19p13.11 (Figure 2A). No widespread genomic inflation was observed as represented by the genomic inflation factor (λ_GC_ = 1.05), shown in the QQ-plot (Figure 2B).

**Figure 2.**
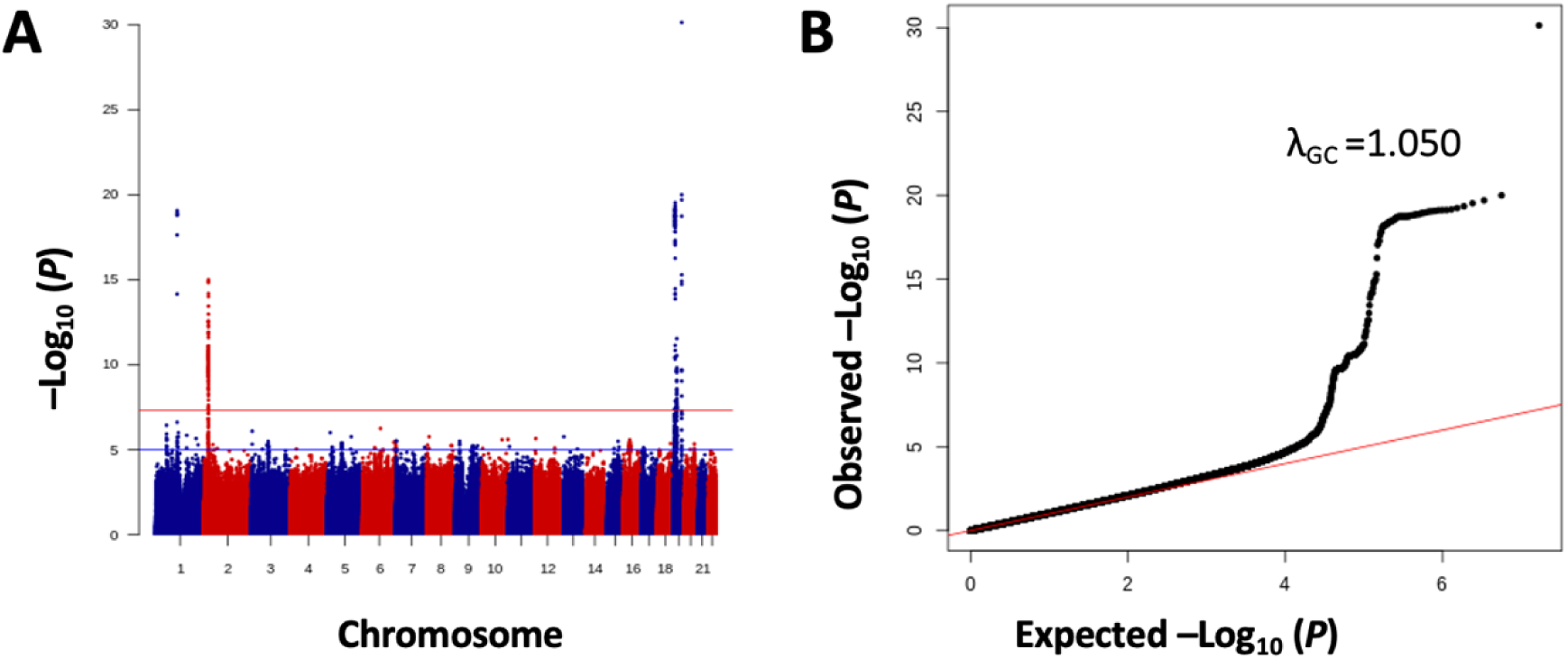
Manhattan and Quantile-Quantile plots from QBB GWAS discovery cohort. **(A)** Manhattan plot represents genetic variants (dots) plotted on x-axis in accordance with chromosome position against their corresponding -log_10_(*P*). The horizontally marked red line indicates the genome-wide significance threshold (*P*=5.0×10^−8^). The horizontal blue line indicates the suggestive significance threshold (*P*=5.0×10^−5^). **(B)** Quantile-Quantile plot shows the expected versus observed -Log_10_ (*P*); λGC is the genomic inflation factor.

GWAS for the discovery dataset consisted of 8,399,151 variants, of which 5 loci (242 variants) reached genome wide significance (*P*<5.0×10^−8^). Meta-analysis of the discovery and replication cohort did not identify any novel LDL-C associated variants (data not shown).

Next, we compared our findings from the GWAS of the discovery dataset with summary statistics of 86,870 genome-wide significant LDL-C associated variants obtained from the Graham *et. al.,* study (10) to assess the level of replication in our cohort. We replicated 8,043 variants (168 loci) at nominal significance (P<0.05) in the QBB dataset (Supplementary Table 2). Comparing the allele frequencies and effect sizes (Beta) of replicated variants to data from the Graham *et. al.,* study showed high correlation of allele frequencies between the two datasets (R^2^= 0.77, Figure 3A) and an overall consistent direction of effect, showing a significant trend for higher effect sizes in QBB cohort (Regression slope = 0.375; P<0.0001; R^2^ = 0.53, Figure 3B).

**Figure 3.**
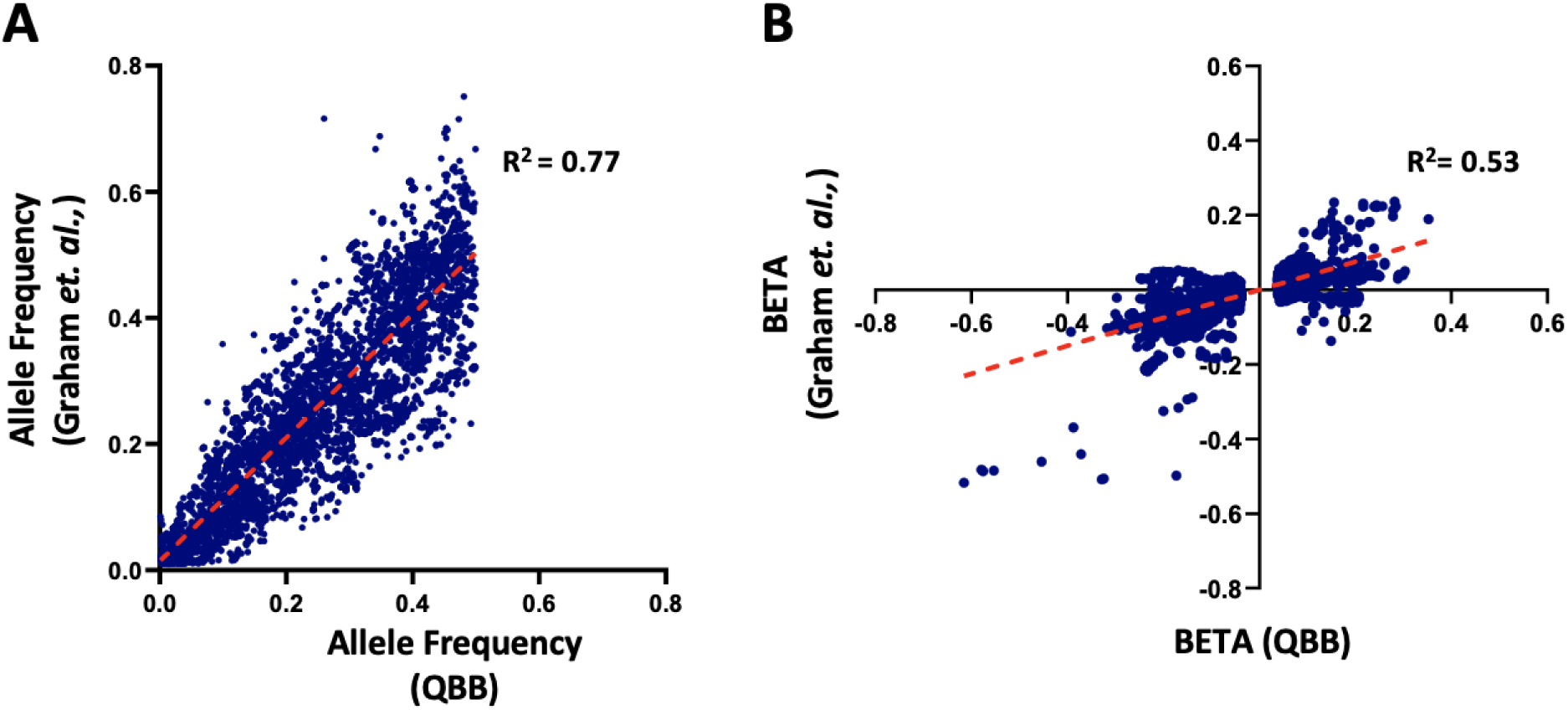
Comparison of allele frequency and effect size for replicated genetic variants. Scatter plots represent the **(A)** Allele frequency and **(B)** Effect size (Beta) comparisons between the QBB discovery cohort and data from the multi-ancestry GWAS study by Graham *et. al.,* R^2^ is the coefficient of determination from correlation analysis. The red dotted line represents the line of best fit from linear regression.

### Multi-ancestry meta-analysis of LDL-C

Next, we performed a MR-MEGA multi-ancestry meta-analysis using the QBB cohort alongside the summary statistics from each single-ancestry GWAS (AFR, EAS, EUR, HIS, SAS) from the Graham *et. al.* study (10). The multi-ancestry meta-analysis yielded 52,621 variants with GC corrected genome-wide significance. Notably, 2,472 of these variants (259 loci) only attained genome-wide significance after the meta-analysis. However, only one of these loci was novel, with no previously reported proxy SNPS in association with LDL-C reported in other studies/databases. The lead variant of our novel locus was rs10939663 (GC corrected *P*=1.25×10^−8^) from the 4p16.1 locus, mapped within the Solute carrier family 2 member 9 *SLC2A9* (Table 1). The regional association plot for the novel locus is shown in Figure 4.

**Table 1.**
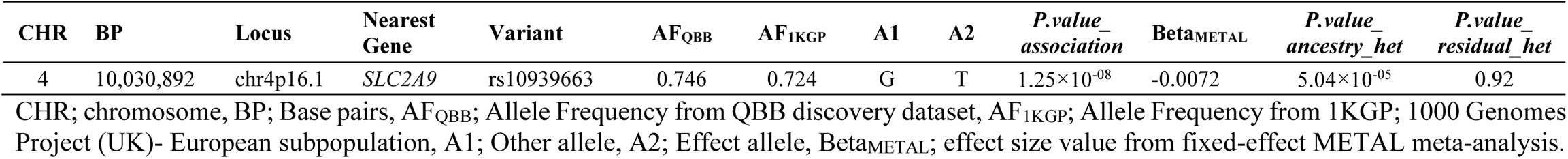
Lead genetic variant from a novel locus identified from multi-ancestry meta-analysis of QBB cohorts with the multi-ancestry summary statistics from Graham *et. al.,* (13).

**Figure 4.**
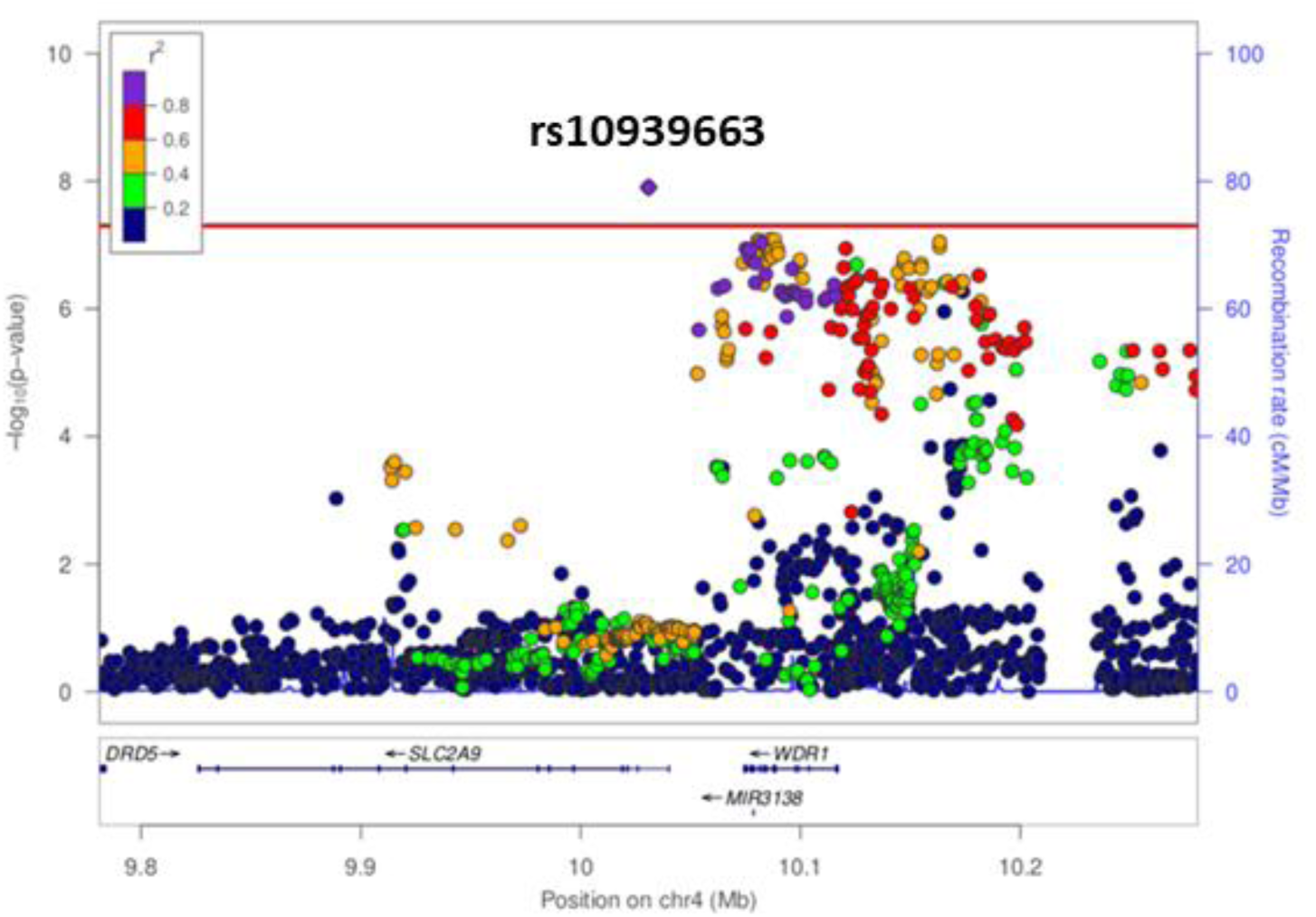
Regional Association plots for novel locus identified from the multi-ancestry meta-analysis for LDL-C. Variants are plotted using the GrCh38 build with x-axis representing the position in the chromosome and the y-axis representing the –log10(*P*-values). rs10939663 located in *SLC2A9.* The linkage disequilibrium (LD) reference from the local population was generated from the QBB data. The color of the plotted dots represents the LD r^2^ value with the lead variants depicted as diamond shaped markers. Recombination rates are also plotted with the corresponding value on the right y-axis. The red line represents -log10 of the genome-wide significance threshold (*P*=5.0×10^−8^).

### Derivation of Qatari-specific PGS panels

The clumping and thresholding methods were used to derive several PGS based on results from LDL-C GWAS of the discovery cohort followed by assessment of their performance in the replication cohort using linear regression models. The PGS panels tested during the optimization stage derived from different clumping LD r^2^ and *P*-values thresholds are listed in Supplementary Table 3. Figure 5 shows the performance values for these PGS panels tested in the replication cohort by adjusting for age, gender, cholesterol treatment and PC1-PC4 in the regression models. Correlation charts for these panels are presented in Supplementary Figures 1-4. The clumping and thresholding method performed better compared to using the thresholding-only method (Figure 5).

**Figure 5.**
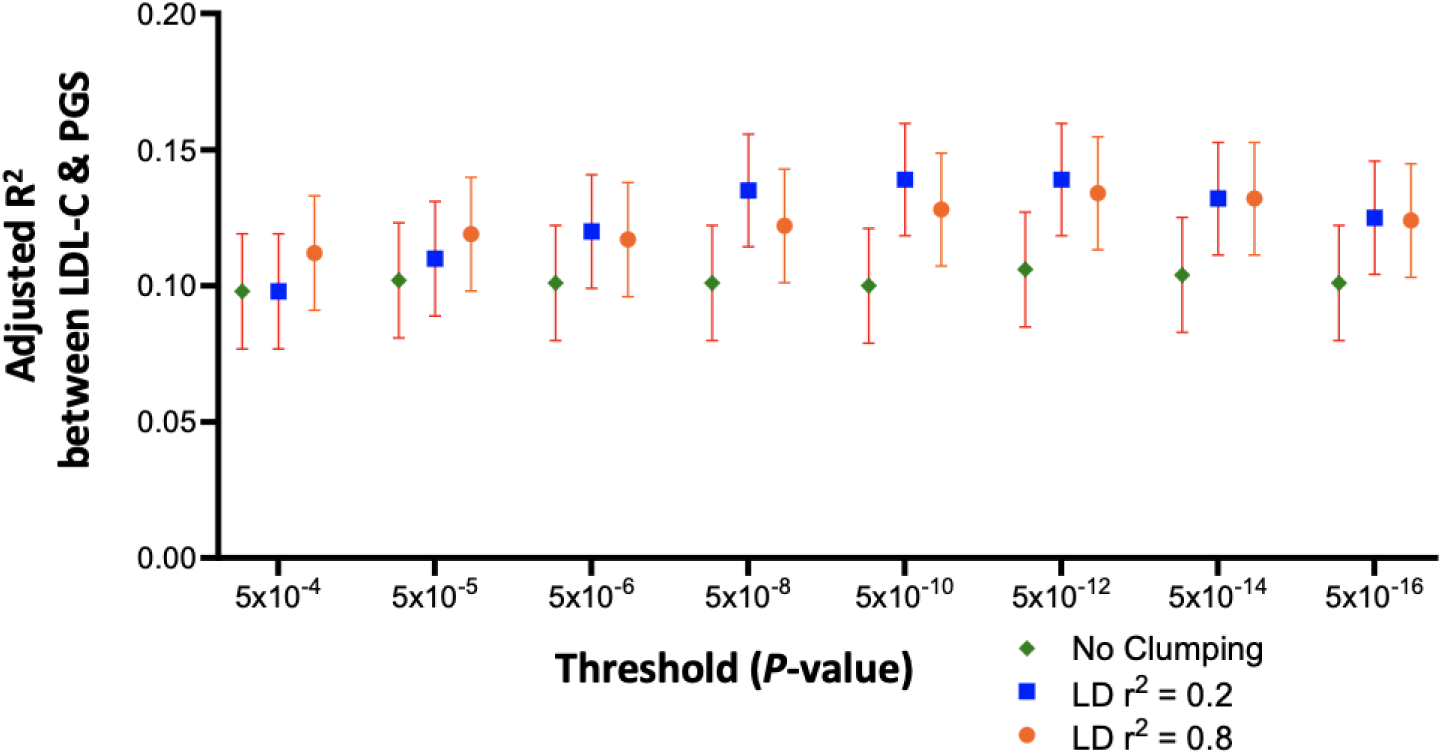
Performance assessment of polygenic score (PGS) panels derived from discovery cohort and tested on the replication cohort. Interleaved scatter plot shows the adjusted R^2^ values from the linear regression models for raw LDL-C values with PGS derived using different-P-value thresholds (x-axis) and clumping LD r^2^ threshold values (0.2 or 0.8). The regression model included the PGS, age, gender, PC1-PC4, and cholesterol treatment as predictors. Data points represent mean adjusted R^2^ with 95% Confidence Intervals (CI). Green diamond datapoints represent PGS derived based on thresholding only while PGS derived based on thresholding and clumping with r^2^ < 0.2 are shown in blue and those with r^2^ < 0.8 are shown in red.

The highest performing PGSs (PGS5_QBB_6SNP, PGS6_QBB_6SNP) were derived from the clumping and thresholding method (LD r^2^=0.2) using a threshold of *P*<5.0×10^−10^ and *P*<5.0×10^−12^ (Table 2). The LDL-C prediction assessment of these best-performing 6-SNP panels resulted in a higher adjusted R^2^ value of 0.14 compared to an 83-SNP PGS derived without clumping using the same *P*-value threshold (adjusted R^2^ value = 0.11; Supplementary Table 4). Moreover, lower LD clumping r^2^ threshold of 0.2 showed a superior performance compared to a higher r^2^ value of 0.8. For example, thresholding at P< 5.0×10^−12^ and clumping at r^2^=0.8 resulted in adjusted R^2^ value of 0.13 compared with 0.14 at LD r^2^=0.2 (Figure 5). The top three performing PGS panels were selected for additional assessment and comparison with other previously derived PGS. PGS5_QBB_6SNP and PGS6_QBB_6SNP showed best performance (adj-R^2^ = 0.139) and were found to contain the same SNPs. PGS5_QBB_6SNP was selected and two additional PGS panels (PGS4_QBB_9SNP; Adj-R^2^ = 0.135 and PGS14_QBB_14SNP; Adj-R^2^ = 0.134) were selected for downstream comparisons (Supplementary Table 4). The 3 best performing panels derived from the LDpred-2 were PGS25_QBB_269K_SNP (adj-R^2^= 0.10), PGS26_QBB_282K_SNP (adj-R^2^= 0.11) and PGS27_QBB_244K_SNP (adj-R^2^= 0.10).

**Table 2.**
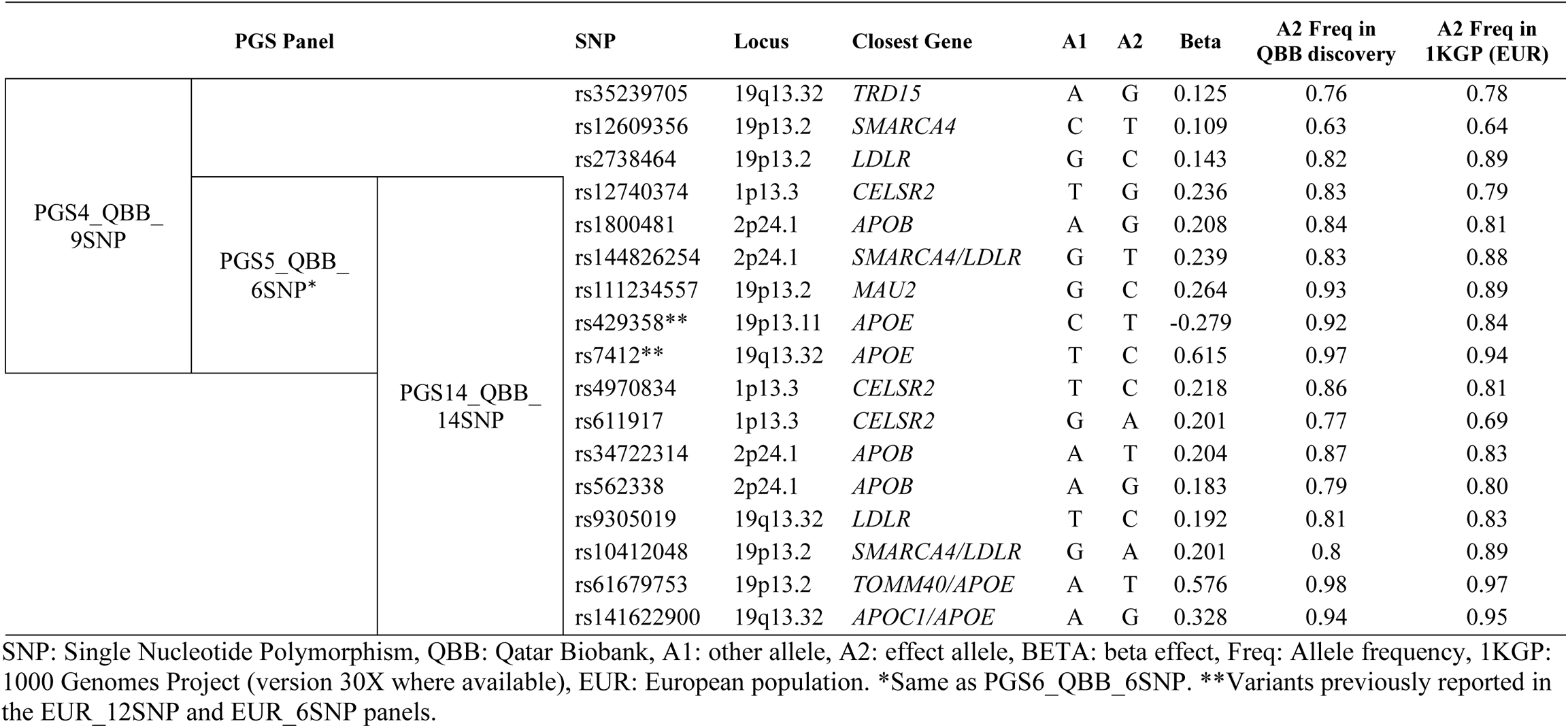
QBB-derived Polygenic score panels for LDL-C.

### Assessment of PGS panel performance

We compared the best performing Qatari-derived LDL-C PGS panels against 9 previously developed LDL-C PGS panels from other ancestries (Supplementary Table 5). These include the first LDL-C derived PGS from European ancestry that included 12 SNPs (EUR_12SNP; PGS000814) (14), a revised version of this PGS that included 6 SNPs (EUR_6SNP) (15), a 36-SNP panel derived from European ancestry (EUR_36SNP; PGS000875) (33), in addition to the best performing LDL-C PGS panels derived from the largest GWAS of European (EUR_1MNSNP; PGS000892, EUR_5427SNP: PGS000893), African (AFR_1MNSNP: PGS000886, AFR_295SNP; PGS000887,) and Multi-ancestry (MULTI_1MNSNP; PGS000888, MULTI_9009SNP; PGS000889) (10). Notably, while all PGS panels included some common SNPs, the total number of SNPs/valid predictors in each PGS panel varied greatly. For instance, 5 of the 6 SNPs from the PGS5_QBB_6SNP occur in common genes as in the EUR_6SNP. The additional variant from the PGS5_QBB_6SNP occurs in *MAU2* (rs111234557), which is not represented in the EUR_6SNP nor the EUR_12SNP panels.

To assess the performance of LDL-C derived PGS panels, we used linear regression models for LDL-C values and included the PGS, age, gender, cholesterol treatment and PCs1-4 as predictors, and as described in Graham et., al. (10). Results showed that the best performing panel was the multiethnic panels (MULTI_1MNSNP; adj-R^2^=0.161 and MULTI_9009SNP panel; adj-R^2^=0.160), followed by the largest European panels (EUR_1MNSNP; adj-R^2^=0.156 and EUR_5427SNP; adj-R^2^=0.141), and the Qatari-derived PGS5_QBB_6SNP (adj-R^2^= 0.139), PGS4_QBB_9SNP (adj-R^2^=0.135) and PGS14_QBB_14SNP (adj-R^2^= 0.134) panels (Figure 6A, Supplementary Table 6 & Supplementary Figure 4). The 3 PGS panels derived using the LDpred-2 method showed lower performance compared to PGS5_QBB_6SNP. Moreover, the AFR_295SNP (adj-R^2^ =0.126) performed better than the 3 European based SNP panels; EUR_6SNP, EUR_12SNP and EUR_36SNP panel. Interestingly, the top performing PGSs showed similar performance when applied to the QBB cohort compared to previously derived PGS applied on their respective ancestries in the associated studies (Figure 6B).

**Figure 6.**
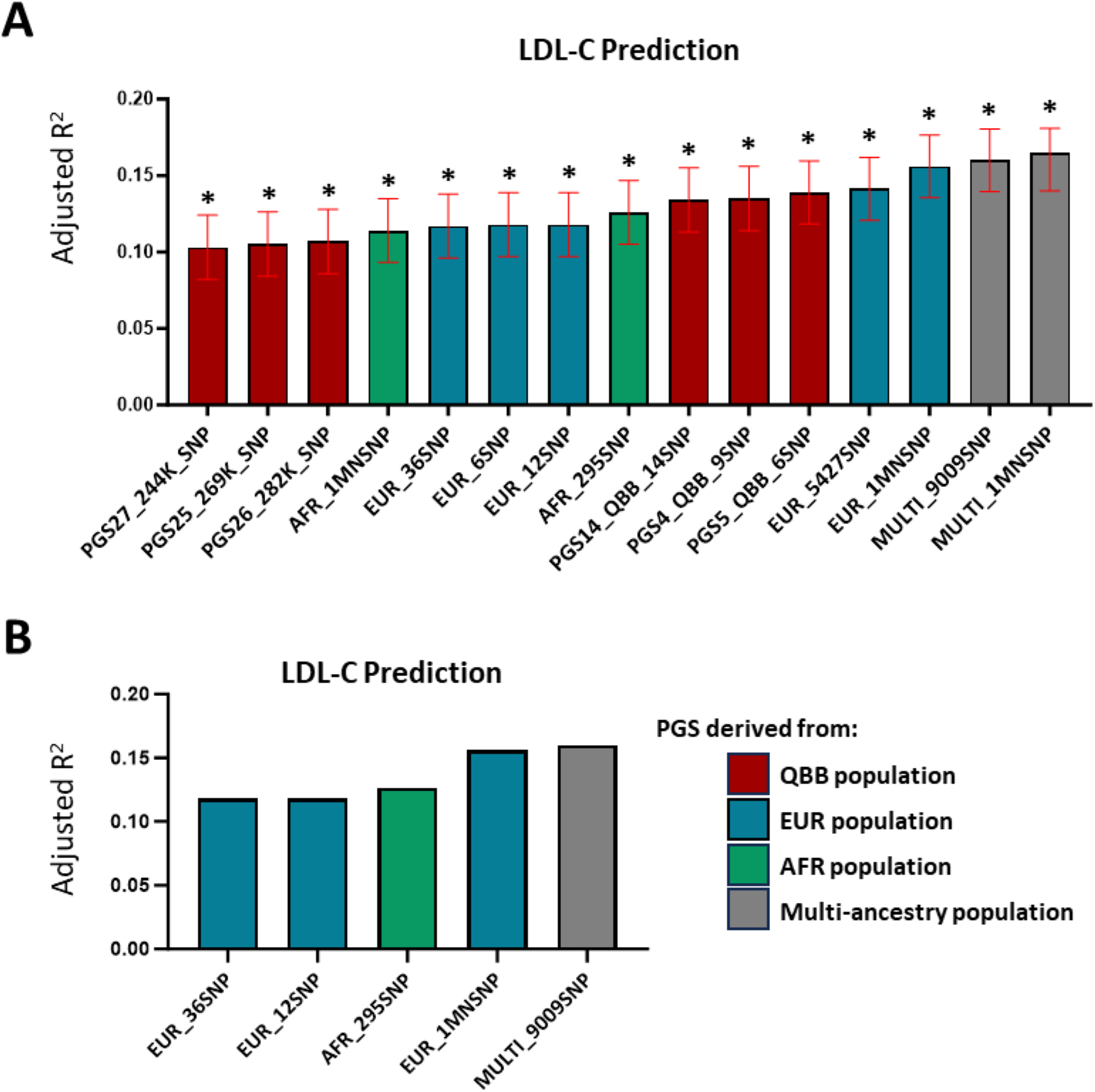
Performance metrics for the polygenic score panels. **(A)** Bar plot shows the adjusted R^2^ values from the performance of the 13 different PGS panels tested on the QBB replication cohort (n=7,762). Performance was assessed using linear regression for raw LDL-C values using PGS, age, gender, PCs1-4, and cholesterol treatment as predictors in the model. **(B)** Bar plot shows the adjusted R^2^ from the LDL-C linear regression models for different PGS panels (n=5); EUR_36SNP (n=4,787) (30), EUR_12SNP (n=3,020) (14) and EUR_1MNSNP (n=389,158) (10) tested on European populations, AFR_295SNP tested on African ancestry (n=6,863) and MULTI_9009SNP tested on multi-ancestry populations (n=461,918) (10). Asterisks (*) represent statistical significance for PGS from regression analysis (*P*<0.05).

## Discussion

The present study represents the largest population-based GWAS to investigate the genomic determinants of LDL-C variation in Middle Eastern and Arab populations. Our GWAS replicated 8,043 variants located in 168 loci in the QBB dataset showing an overall consistent direction of effect indicating a significant trend for higher effect sizes in QBB cohort. The observed variations in effect size estimates of individual variants based on pairwise comparisons between ancestries could indicate diverse patterns of linkage disequilibrium (LD); with the underlying causal variant, or a potential interaction with an environmental risk factor that exhibits varying prevalence across different ancestral backgrounds and/or geographical regions.

Moreover, the multi-ancestry meta-analysis with the largest GWAS of LDL-C disclosed a novel locus in a gene that has not been previously associated with LDL-C concentration in blood. Our findings reflect the importance of inclusion of Middle eastern ancestral populations in GWAS since this novel locus exclusively reached genome-wide significance following the multi-ancestry meta-analysis. The identified LDL-C-associated novel locus was located in the intronic region of *SLC2A9*. This variant was initially below the threshold for genome-wide significance in its original 7 cohorts (i.e., QBB discovery, QBB replication and 5 Graham et al. single ancestry cohorts) but when combined, it showed genome-wide significant association with LDL-C after genomic control correction. This gene has not been previously associated with LDL-C levels in the GWAS catalog (9), Phenoscanner (35) or the Common Metabolic Disease (CMD) Knowledge Portal (29).

The *Solute Carrier Family 2 Member 9* (*SLC2A9*) gene encodes a member of the SLC2A facilitative glucose transporter family (GLUT9), which is involved in the transportation of glucose and uric acid (36). Variants of *SLC2A9* are strongly associated with serum uric acid levels and gout (37, 38) in multiple ancestry individuals (39, 40). Importantly, while previous GWAS have predominantly associated variants in *SLC2A9* with metabolite levels, hypouricemia is also associated with serum lipid profiles (41, 42). Our findings present a variant in *SLC2A9* as a novel LDL-C-associated gene and its involvement in molecular pathways involved in cardiovascular disease progression and outcomes may be explored further.

Combined, while our identified novel LDL-C-associated gene has been previously implicated with different pathological conditions, the impact of its genetic deviation on gene regulatory network or molecular basis of LDL-C regulation, associated risk of CVD development & progression warrant further investigations.

LDL-C levels can serve as early risk predictors of CVD and associated complications. PGS for LDL-C have been extensively explored in other populations to assess the genomic risk of hypercholesteremia and associated CVD risk, with varying performance and limited clinical utilization (43). While the genes included in our QBB PGS panels have also been previously associated with LDL-C levels, the specific set of variants has not been previously identified as the optimal representative panel to accurately predict LDL-C levels in other ancestries. For instance, the PGS5_QBB_6SNP panel, 4 out of the 6 variants were specific to the Qatari population compared to the EUR_6/12SNP panel demonstrating the variation in genetic architecture between the two populations. Additionally, our identified variants occurred in genes similar to the EUR_6/12SNP panels (*LDLR, CELSR2, APOB,* and *APOE*) (14, 15) however, one variant in *MAU2* was only present in the Qatari PGS panel.

In concordance with the Graham *et. al.,* study, which reported that the MULTI_9009SNP PGS panel showed equivalent or better performance across most ancestry groups compared to the ancestry-specific PRS (10), this multi-ancestry derived panel also showed the best performance for LDL-C prediction in our Qatari cohort. Polygenic scores derived from multiethnic ancestries can capture a wider range of genetic variations related to LDL-C and provide more accurate predictions for LDL-C trait. However, the performance metrics of the 3 Qatari-specific PRS were slightly lower (0.021 to 0.026 lower adj-R^2^ value) than the MULTI-9009SNP PGS. Importantly, our QBB PGS panels comprised of considerably fewer variants than the PGS panels from other ethnicities (6 variants in the Qatari panel compared to >1 million or 9,009 variants in the multi-ancestry PGS panels), indicating the potent association of these sets of variants in LDL-C prediction within the Qatari population. Moreover, fewer SNPs in our PGS panels also support more feasible potential clinical utilization. Additionally, our PGS panel was derived from whole genome sequence data providing more comprehensive coverage of genetic variations compared to genotyping arrays and imputation methods used in most previous GWAS. Of note, the EUR_12SNP and EUR_36SNP PGS showed stronger performances in predicting LDL-C variance in our study compared to their original studies. However, the analysis models used in the EUR_12SNP and EUR_36SNP studies were different from the model parameters used in our study, hindering accurate comparisons of performance metrics between the studies. The LDpred-2 models from our dataset did not perform as strongly as our C+T models possibly because of our small cohort size or because they need to be further optimized for our dataset (e.g. using a larger set of variants rather than restricting to common variants with HapMap). The benefits of this method are more obvious when dealing with larger sample sizes (44).

Although the findings from this study provide new insights into the genetic architecture of LDL-C, our study has some limitations. While the present study is the largest GWAS of LDL-C from a Middle Eastern population, it is relatively modest compared to recent published GWAS studies of LDL-C from European ancestry. However, in agreement with the findings of Graham *et. al.,* the inclusion of individuals from diverse ancestries, such as the underrepresented Middle Eastern population, in meta-analysis leads to enhanced discoveries of novel variants and genomic regions for LDL-C as we demonstrated herein. Further larger studies from Middle Eastern populations will be required to capture more genetic variations from this ancestry.

## Conclusion

Our study utilized population-based GWAS using WGS to identify variants that are associated with LDL-C levels within the Qatari population. We also performed a multi-ancestry meta-analysis with the largest LDL-C meta-analysis previously reported by the GLGC consortium (10). We replicated 168 genetic loci between QBB and Graham *et. al.,* datasets and their allele frequencies and beta-effect values were broadly concordant. Importantly, we identified 1 novel LDL-C-associated locus in the multi-ancestry meta-analysis, indicating the importance of increasing ancestral diversity in GWAS. Additional investigations to decipher the molecular and pathophysiological roles of our identified variants in affecting LDL-C concentration in blood would facilitate identification of therapeutic targets. The multi-ancestry derived LDL-C PGS (MULTI_1MNSNP) performed best at predicting LDL-C in our cohort. In addition, we developed 3 Qatari-based LDL-C PGS panels that performed reasonably well in predicting LDL-C levels. LDL-C level prediction has limited clinical utility since LDL-C levels are routinely tested in clinical practice. However, identification of genetic variations and their effects on molecular pathways leading to variation in LDL-C levels has potential for disclosing therapeutic targets. Moreover, development of PGS to reflect genetic predisposition to elevated LDL-C levels can aid in mitigating risk via administration of medical or lifestyle interventions.

## Author contributions

U.I.U: Data Curation, Formal Analysis, Investigation and Writing—original draft. S.M.T.: Investigation, Writing—original draft and Visualization. Y.A.S: Data curation, Investigation, Methodology. S.M./M.S.AH./E.U/K.K/A.EM/A.J./M.S/N.Q/J.M.S: Resources. O.M.E.A.: Conceptualization, Funding Acquisition, Formal Analysis, Supervision, Investigation and Writing-review and editing.

## Acknowledgements

We are grateful to the Qatar Biobank (QBB) and the Qatar Genome Program (QGP) for providing access to the genomic and phenotypic data. We are also thankful to all participants of this study. This study was supported by a grant from the Qatar National Research Fund (QNRF) awarded to OMEA (PPM 03-0324-190038).

## Data availability

The data analyzed in this study are subject to the following licenses/restrictions: the raw WGS data from the QBB are protected and are not available for deposition into public databases due to data privacy laws. Access to QBB/QGP phenotype and whole-genome sequence data can be obtained through an ISO-certified protocol, which involves submitting a project request at https://www.qatarbiobank.org.qa/research/how-apply, subject to approval by the Institutional Review Board of the QBB. Requests to access these datasets should be directed to https://www.qatarbiobank.org.qa/research/how-apply.

