## Supplementary Figures for "Identification of a Novel *SLC2A9* Gene Association with LDL-C levels and Evaluation of Polygenic Scores in a Multi-Ancestry Genome Wide Association Study"

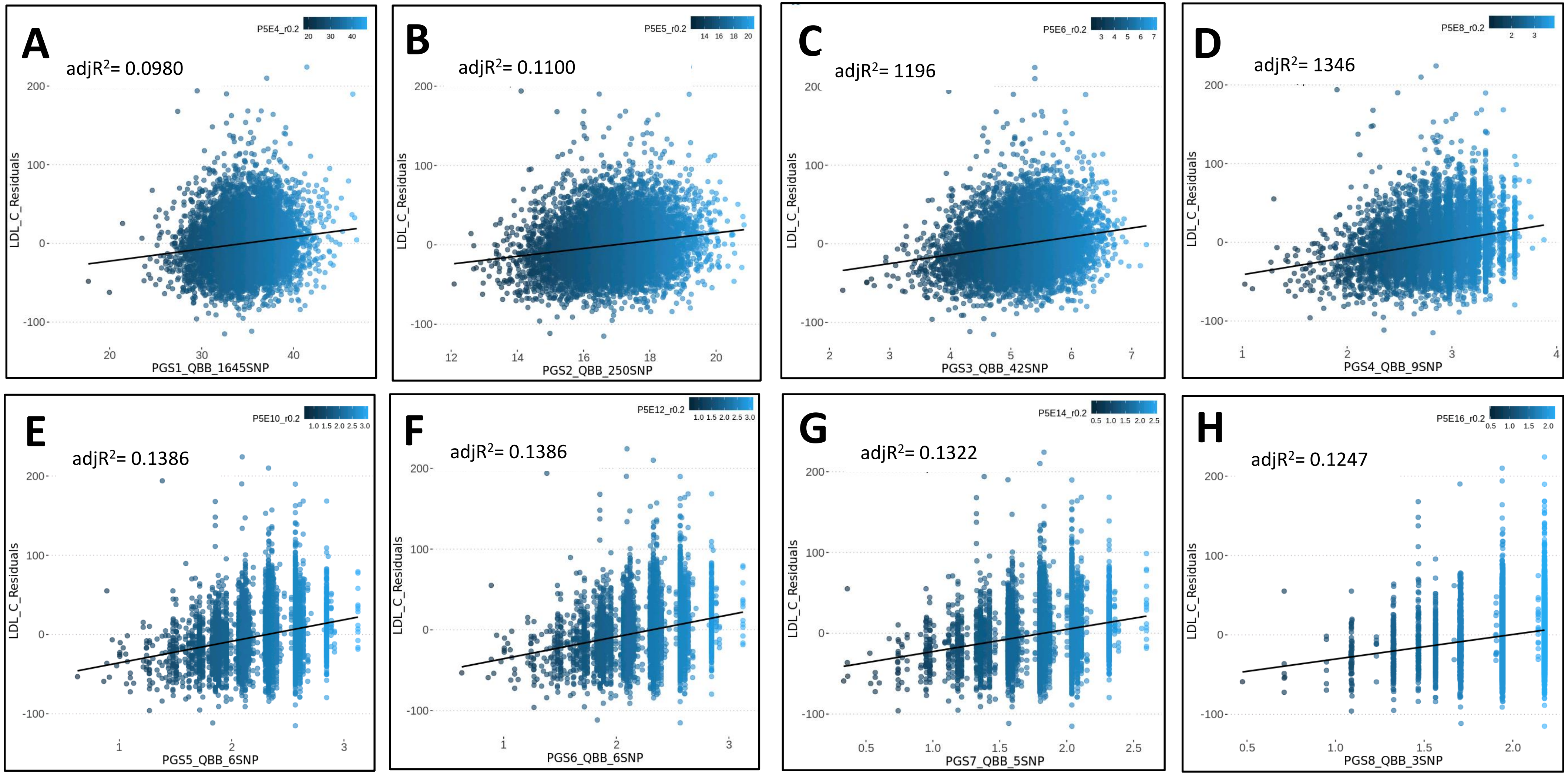


**Supplementary Figure 1. Correlation plots of LDL-C with PGS1-PGS8 based on linear regression models.** Scatter plots show the correlation between PGS and LDL-C residuals for **(A)** PGS1-QBB_1645SNP, **(B)** PGS2_QBB_250SNP, **(C)** PGS3_QBB_42SNP, **(D)** PGS4_QBB_9SNP, **(E)** PGS5_QBB_6SNP, **(F)** PGS6_QBB_6SNP, **(G)** PGS7_QBB_5SNP, **(H)** PGS8_QBB_3SNP. LDL-C values were adjusted for age, gender, PCs1-4, and cholesterol treatment and residuals were calculated. PGS panels were generated using different *P*-values and clumping + thresholding techniques at LD-clump-$r^2<0.2$. The colour intensity represents increasing PGS score (the lighter the colour the higher the score)

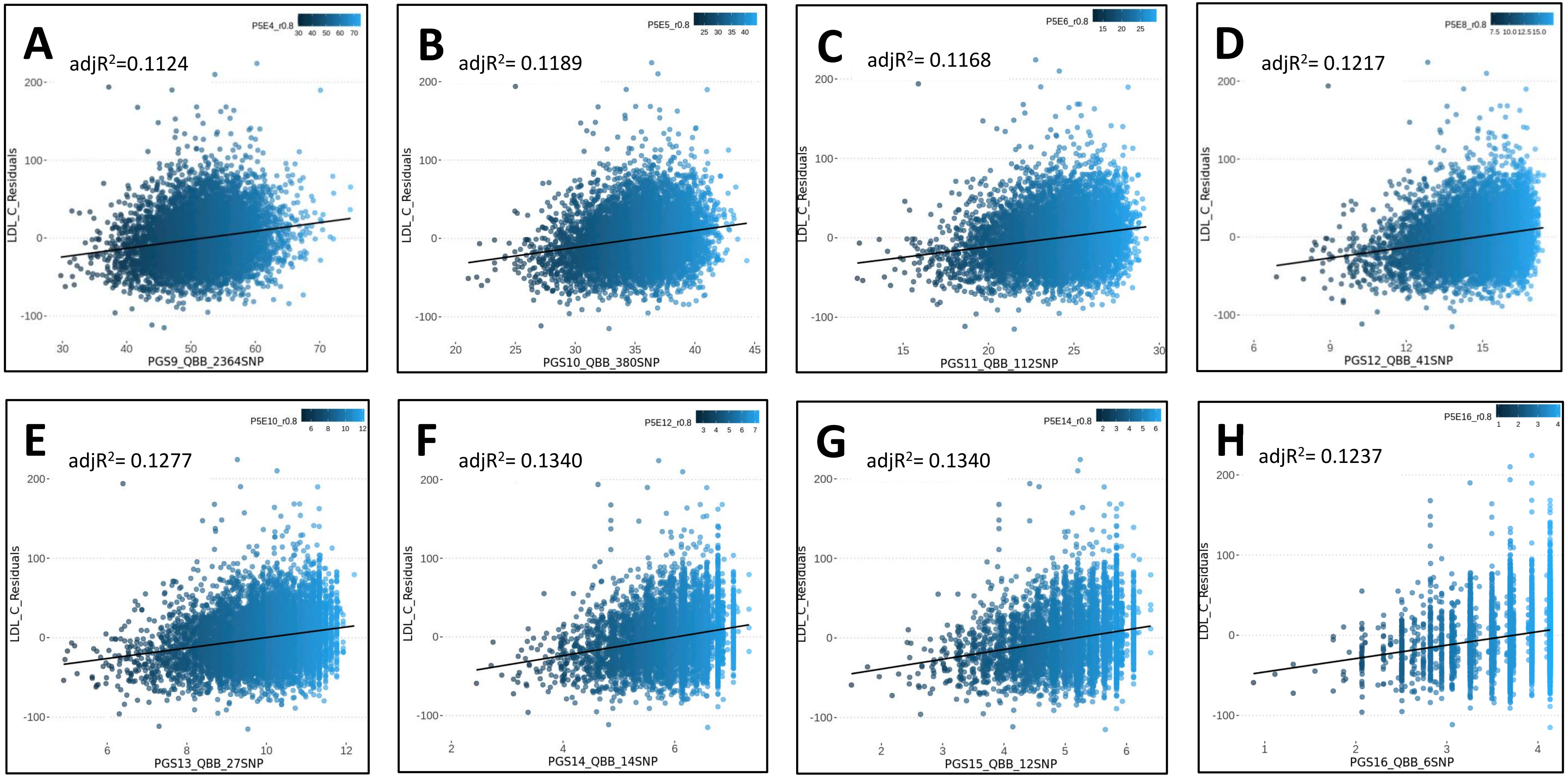


**Supplementary Figure 2. Correlation plots of LDL-C with PGS9-PGS16 based on linear regression models.** Scatter plots show the correlation between PGS and LDL-C residuals for **A)** PGS9_QBB_2364SNP**, (B)** PGS10_QBB_380SNP**, (C)** PGS11_QBB_112SNP**, (D)** PGS12_QBB_41SNP**, (E)** PGS13_QBB_27SNP**, (F)** PGS14_QBB_14SNP**, (G)** PGS15_QBB_12SNP**, (H)** PGS16_QBB_6SNP**.** LDL-C values were adjusted for age, gender, PCs1-4, and cholesterol treatment and residuals were calculated. PGS panels were generated using different *P*-values and clumping + thresholding techniques at LD-clump-$r^2$<0.8. The colour intensity represents increasing PGS score (the lighter the colour the higher the score).

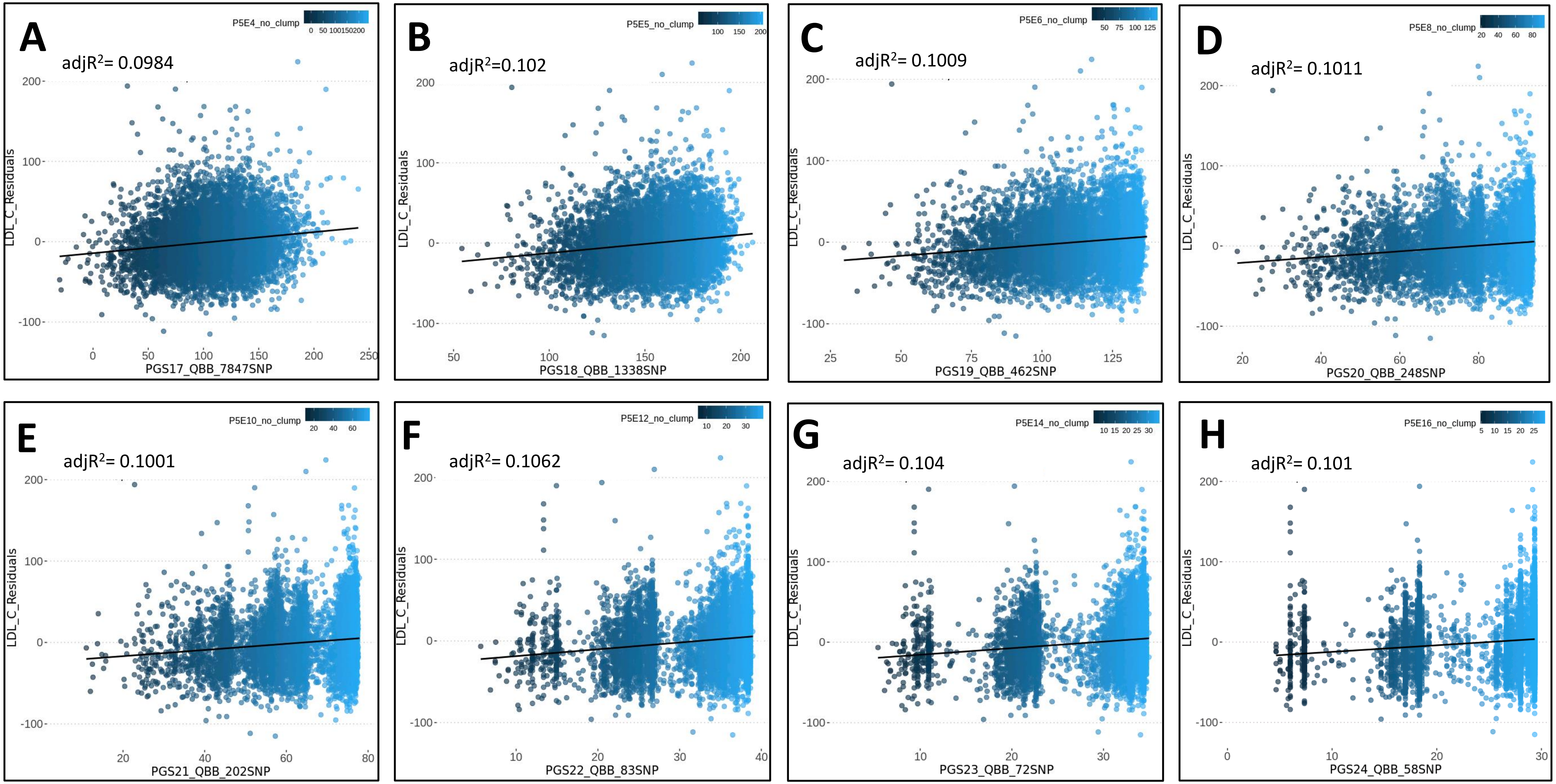


**Supplementary Figure 3. Correlation plots of LDL-C residuals with PGS17-PG24 based on linear regression models.** Scatter plots show the correlation between PGS and LDL-C residuals for **(A)** PGS17_QBB_7847SNP**, (B)** PGS18_QBB_1338SNP**, (C)** PGS19_QBB_462SNP**, (D)** PGS20_QBB_248SNP**, (E)** PGS21_QBB_202SNP**, (F)** PGS22_QBB_83SNP**, (G)** PGS23_QBB_72SNP**, (H)** PGS24_QBB_58SNP. LDL-C values were adjusted for age, gender, PCs1-4, and cholesterol treatment and residuals were calculated. PGS panels were generated using different *P*-values thresholding only. The colour intensity represents increasing PGS score (the lighter the colour the higher the score)

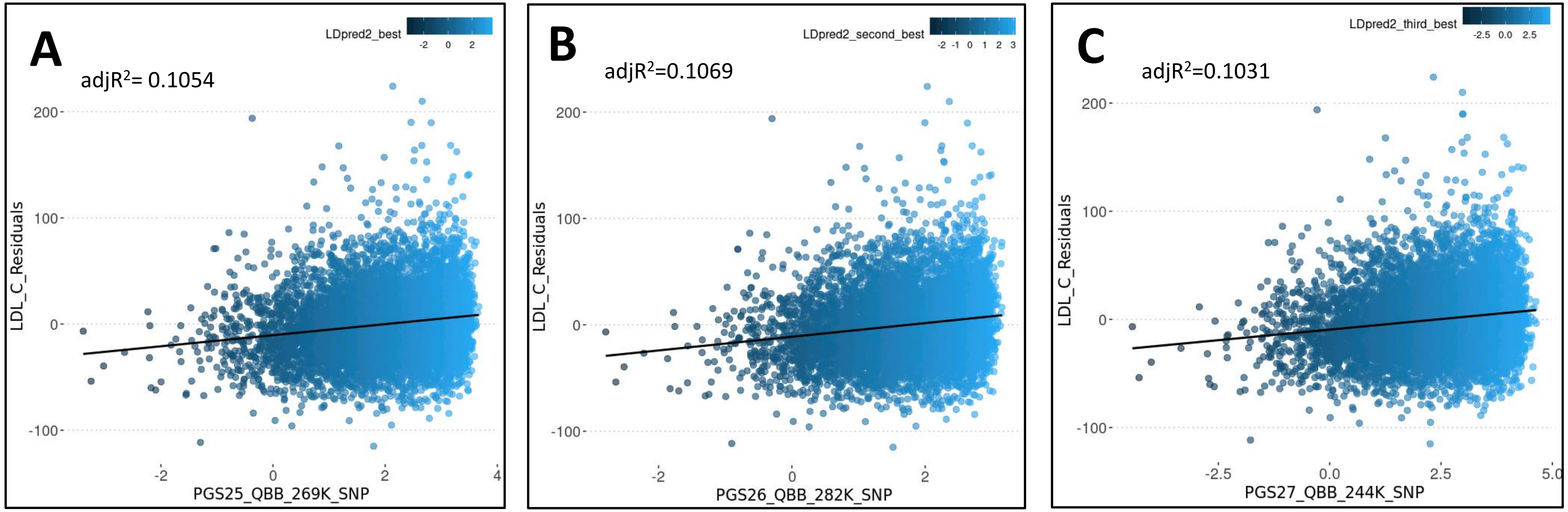


**Supplementary Figure 4. Correlation plots for LDL-C residuals with PGS25-PGS27 based on linear regression models.** The best performing QBB panels (n=3) derived from LDPred2. Scoring was performed on the QBB replication cohort (n=7762) without an allele frequency cut-off. Scatter plots show correlation between PGS and LDL-C residuals for **(A)** PGS25_QBB_269K_SNP, **(B)** PGS26_QBB_282K_SNP, **(C)** PGS27_QBB_244K_SNP. LDL-C values were adjusted for age, gender, PCs1-4, and cholesterol treatment and residuals were calculated. The colour intensity represents increasing PGS score (the lighter the colour the higher the score).

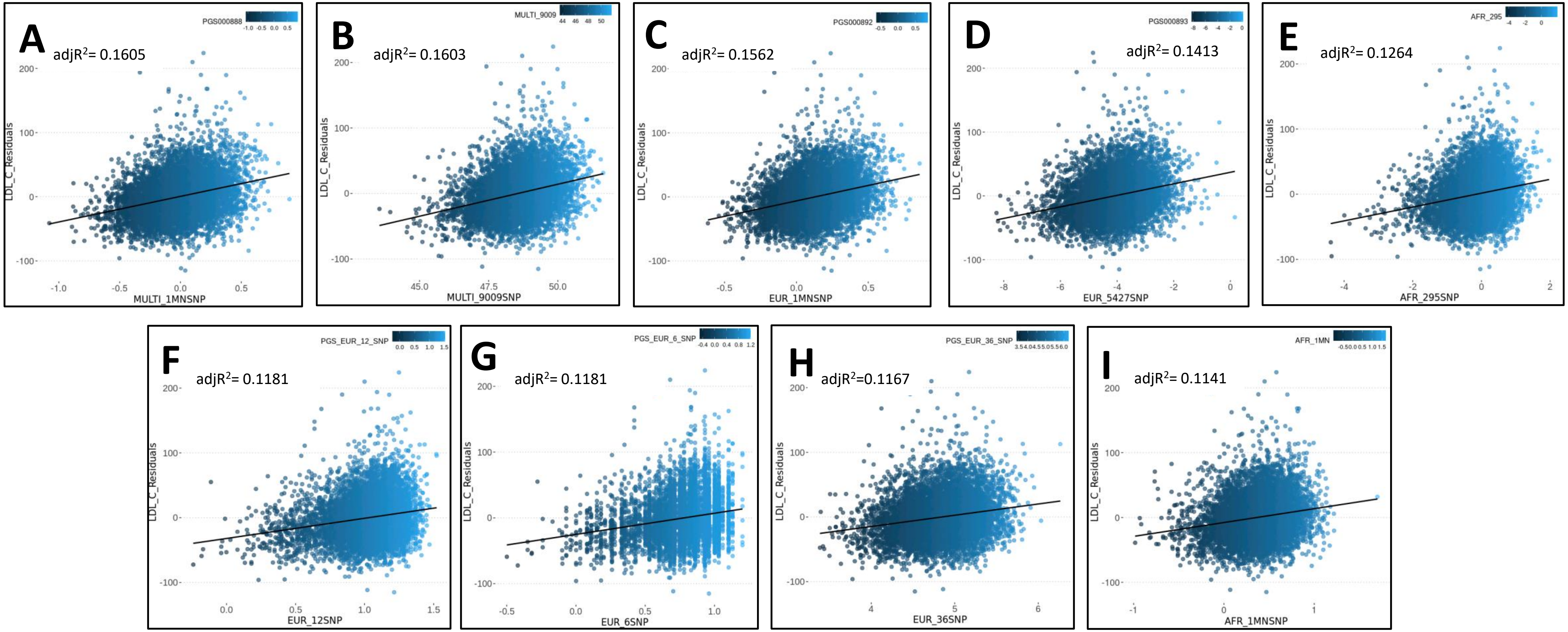


**Supplementary Figure 5. Correlation plots for LDL-C with previously derived PGSs based on linear regression models.** Scoring was performed on the QBB replication cohort (n=7762) without an allele frequency cut-off. Scatter plots show correlation between PGS and LDL-C residuals for **(A)** MULTI_1MNSNP **(B)** MULTI_9009SNP **(C)** EUR_1MNSNP **(D)** EUR_5427SNP **(E)** AFR_295SNP **(F)** EUR_12SNP **(G)** EUR_6SNP **(H)** EUR_36SNP **(I)** AFR_1MNSNP. LDL-C values were adjusted for age, gender, PCs1-4, and cholesterol treatment and residuals were calculated. The colour intensity represents increasing PGS score (the lighter the colour the higher the score).
