## Supplementary Table 1 for "Identification of a Novel *SLC2A9* Gene Association with LDL-C levels and Evaluation of Polygenic Scores in a Multi-Ancestry Genome Wide Association Study"

**Supplementary Table 1.** Clinical characteristics of study cohort.

| Cohort | Discovery | Replication |
| --- | --- | --- |
| **Number of subjects** | 5,939 | 7,762 |
| **Age (mean ± SD) *** | 39.7±12.8 | 40.4±13.4 |
| **Male** | 2,534 | 3,500 |
| **Female** | 3,355 | 4,262 |
| **BMI[I] (kg/m²) *** | 29.4±6.1 | 29.7±6.2 |
| **LDL-C (mg/dL) *** | 114.9±34.6 | 111.9 ±34.5 |

Quantitative variables are presented as mean ± standard deviation.
*Statistically significant ($P < 0.05$). [I]BMI: body mass index
