## Supplementary Table 3 for "Identification of a Novel *SLC2A9* Gene Association with LDL-C levels and Evaluation of Polygenic Scores in a Multi-Ancestry Genome Wide Association Study"

**Supplementary Table 3**. Polygenic Risk Score Panels derived from the QBB discovery GWAS.

| | **Clumping and thresholding only*** | | | | **Thresholding only** | | **Bayesian Shrinkage**** | |
|---|---|---|---|---|---|---|---|---|
| | **LD clump $r^2$=0.2*** | | **LD clump $r^2$=0.8*** | | | | | |
| ***P* value threshold** | **PGS Name** | **No. of clumps (variants)** | **PGS Name** | **No. of clumps (variants)** | **PGS Name** | **No. of variants** | **PGS Name** | **No. of variants** |
| $5.0 \times 10^{-4}$ | PGS1_QBB_1645SNP | 1645 (7847) | PGS9_QBB_2364SNP | 2364 (7847) | PGS17_QBB_7302SNP | 7847 | PGS25_QBB_269KSNP | 269,694 |
| $5.0 \times 10^{-5}$ | PGS2_QBB_250SNP | 250 (1338) | PGS10_QBB_380SNP | 380 (1338) | PGS18_QBB_1352SNP | 1338 | PGS26_QBB_282KSNP | 282,998 |
| $5.0 \times 10^{-6}$ | PGS3_QBB_42SNP | 42 (462) | PGS11_QBB_112SNP | 112 (462) | PGS19_QBB_449SNP | 462 | PGS27_QBB_244KSNP | 244,787 |
| $5.0 \times 10^{-8}$ | PGS4_QBB_9SNP | 9 (248) | PGS12_QBB_41SNP | 41 (248) | PGS20_QBB_230SNP | 248 | | |
| $5.0 \times 10^{-10}$ | PGS5_QBB_6SNP | 6 (202) | PGS13_QBB_27SNP | 27 (202) | PGS21_QBB_195SNP | 202 | | |
| $5.0 \times 10^{-12}$ | PGS6_QBB_6SNP | 6 (83) | PGS14_QBB_14SNP | 14 (83) | PGS22_QBB_85SNP | 83 | | |
| $5.0 \times 10^{-14}$ | PGS7_QBB_5SNP | 5 (72) | PGS15_QBB_12SNP | 12 (72) | PGS23_QBB_72SNP | 72 | | |
| $5.0 \times 10^{-16}$ | PGS8_QBB_3SNP | 3(58) | PGS16_QBB_6SNP | 6 (58) | PGS24_QBB_58SNP | 58 | | |

QBB: Qatar Biobank, SNP: Single nucleotide polymorphisms, LD; Linkage disequilibrium, $r^2$; Squared correlation.
