## Supplementary Table 4 for "Identification of a Novel *SLC2A9* Gene Association with LDL-C levels and Evaluation of Polygenic Scores in a Multi-Ancestry Genome Wide Association Study"

**Supplementary Table 4:** Predictive performance of QBB polygenic risk score panels.

| Clumping and Thresholding* | | | | Thresholding Only | | Bayesian Shrinkage** | |
|---|---|---|---|---|---|---|---|
| LD-clump-$r^2$ =0.2 | | LD-clump-$r^2$ =0.8 | | | | | |
| **PGS Name** | **Adj- $R^2$ [95%CI]** | **PGS Name** | **Adj- $R^2$ [95%CI]** | **PGS Name** | **Adj-$R^2$ [95%CI]** | **PGS Name** | **Adj-$R^2$ [95%CI]** |
| PGS1_QBB_1645SNP | 0.098 [0.077-0.119] | PGS9_QBB_2364SNP | 0.112 [0.091-0.133] | PGS17_QBB_7847SNP | 0.098 [0.077-0.119] | PGS25_QBB_269K_SNP | 0.1054 [0.084-0.127] |
| PGS2_QBB_250SNP | 0.110[0.089-0.131] | PGS10_QBB_380SNP | 0.119 [0.098-0.140] | PGS18_QBB_1338SNP | 0.102 [0.081-0.123] | PGS26_QBB_282K_SNP | 0.1069 [0.086-0.128] |
| PGS3_QBB_42SNP | 0.120 [0.099-0.141] | PGS11_QBB_112SNP | 0.117 [0.096-0.138] | PGS19_QBB_462SNP | 0.101 [0.080-0.122] | PGS27_QBB_244K_SNP | 0.1031 [0.082-0.124] |
| PGS4_QBB_9SNP | 0.135 [0.114-0.156] | PGS12_QBB_41SNP | 0.122 [0.101-0.143] | PGS20_QBB_248SNP | 0.101 [0.080-0.122] | | |
| PGS5_QBB_6SNP | 0.139 [0.118-0.160] | PGS13_QBB_27SNP | 0.128 [0.107-0.149] | PGS21_QBB_202SNP | 0.100 [0.079-0.121] | | |
| PGS6_QBB_6SNP | 0.139 [0.118-0.160] | PGS14_QBB_14SNP | 0.134 [0.113-0.155] | PGS22_QBB_83SNP | 0.106 [0.085-0.127] | | |
| PGS7_QBB_5SNP | 0.132 [0.111-0.153] | PGS15_QBB_12SNP | 0.132 [0.111-0.153] | PGS23_QBB_72SNP | 0.104 [0.083-0.125] | | |
| PGS8_QBB_3SNP | 0.125 [0.104-0.146] | PGS16_QBB_6SNP | 0.124 [0.103-0.145] | PGS24_QBB_58SNP | 0.101 [0.080-0.122] | | |

QBB: Qatar Biobank, LD; Linkage disequilibrium, $r^2$; Squared correlation, Adj-$R^2$; adjusted $R^2$. The 95% confidence intervals (CI) are presented within brackets.
