## Supplementary Table 5 for "Identification of a Novel *SLC2A9* Gene Association with LDL-C levels and Evaluation of Polygenic Scores in a Multi-Ancestry Genome Wide Association Study"

**Supplementary Table 5.** Summary of Polygenic Risk scores compared in this study.

| PGS | SNPs | Valid Predictors (QBB) | PGS Name used in this study | Ethnicity | Associated Study |
|---|---|---|---|---|---|
| PGS4_QBB_9SNP | 9 | 9 | PGS4_QBB_9SNP | Arab-Qatari | Current Study |
| PGS5_QBB_6SNP | 6 | 6 | PGS5_QBB_6SNP | Arab-Qatari | |
| PGS14_QBB_14SNP | 14 | 14 | PGS14_QBB_14SNP | Arab-Qatari | |
| PGS25_269K_SNP | 269,694 | 269,475 | PGS25_269K_SNP | Arab-Qatari | |
| PGS26_282K_SNP | 282,998 | 282,797 | PGS26_282K_SNP | Arab-Qatari | |
| PGS27_244K_SNP | 244,787 | 244,614 | PGS27_244K_SNP | Arab-Qatari | |
| PGS000886 | 1,222,318 | 1,023,875 | AFR_1MNSNP | African | Graham SE *et. al.*, Nature (2021) |
| PGS000887 | 295 | 254 | AFR_295SNP | African | |
| PGS000888 | 1,239,184 | 1,033,528 | MULTI_1MNSNP | Multi-Ethnic | |
| PGS000889 | 9,009 | 7,279 | MULTI_9009SNP | Multi-Ethnic | |
| PGS000892 | 1,119,212 | 933,604 | EUR_1MNSNP | European | |
| PGS000893 | 5,427 | 4,283 | EUR_5427SNP | European | |
| Refined-PGS000814 | 6 | 6 | EUR_6SNP | European | Futema *et. al.*, Clin Chem (2015) |
| PGS000814 | 12 | 11 | EUR_12SNP | European | Talmud PJ *et. al.*, Lancet (2013) |
| PGS000875 | 36 | 31 | EUR_36SNP | European | Leal LG *et. al.*, Mol Genet Genomic Med (2020) |

PGS; Polygenic Risk Score, SNP: Single Nucleotide Polymorphisms, SNPs: number of SNPs in the original panel. Valid predictors: number of variants from each panel present and scored in the QBB data without allele frequency filtration.
