## Supplementary Table 6 for "Identification of a Novel *SLC2A9* Gene Association with LDL-C levels and Evaluation of Polygenic Scores in a Multi-Ancestry Genome Wide Association Study"

**Supplementary Table 6**: Best performing QBB-derived PGS panels compared to previously derived PGS panels for LDL-C levels prediction.

| Polygenic Score Panel (reference) | PGS Name used in this study | Linear Regression Model (LDL-C) in QBB* | Comparison with other studies for LDL-C (Adjusted $R^2$) |
|---|---|---|---|
| | | Adjusted $R^2$ [95%CI] | |
| **PGS4_QBB_9SNP** | PGS4_QBB_9SNP | 0.135 [0.114-0.156] | - |
| **PGS5_QBB_6SNP** | PGS5_QBB_6SNP | 0.139 [0.118-0.160] | - |
| **PGS14_QBB_14SNP** | PGS14_QBB_9SNP | 0.134 [0.113-0.155] | - |
| **PGS25_269K_SNP** | PGS25_269K_SNP | 0.105 [0.084-0.127] | - |
| **PGS26_282K_SNP** | PGS26_282K_SNP | 0.107 [0.086-0.128] | - |
| **PGS27_244K_SNP** | PGS27_244K_SNP | 0.103 [0.082-0.124] | - |
| **Refined_PGS000814** (15) | EUR_6SNP | 0.118 [0.097-0.139] | - |
| **PGS000814** (14) | EUR_12SNP | 0.118 [0.097-0.139] | 0.110 (Age + Gender + BMI + Cholesterol treatment + T2D status + Smoking status + BP) |
| **PGS000875** (31) | EUR_36SNP | 0.117 [0.096-0.138] | 0.08 (Age + Gender + BMI + PC1,2) |
| **PGS000892** (10) | EUR_1MNSNP | 0.156 [0.136-0.176] | 0.182 (Treatment corrected LDL-C~ PGS + Age + Gender + PC1-4 + Batch) |
| **PGS000893** (10) | EUR_5427SNP | 0.141[0.121-0.162] | 0.176 (Treatment corrected LDL-C ~ PGS + Age + Gender +PC1-4 +Batch) |
| **PGS000896** (10) | AFR_1MNSNP | 0.114[0.093-0.135] | 0.112 (Treatment corrected LDL-C ~ PGS + Age + Gender +PC1-4 +Batch) |
| **PGS000887** (10) | AFR_295SNP | 0.126 [0.105-0.147] | 0.175 (Treatment corrected LDL-C~ PGS + Age + Gender + PC1-4 + Batch) |
| **PGS000888** (10) | MULTI_1MNSNP | 0.161[0.140-0.181] | 0.175 (Treatment corrected LDL-C ~ PGS + Age + Gender +PC1-4 +Batch) |
| **PGS000889** (10) | MULTI_9009SNP | 0.160 [0.140-0.180] | 0.182 (Treatment corrected LDL-C~ PGS + Age + Gender + PC1-4 + Batch) |

*All linear regression models tested in QBB cohort had a *P*-value < $2\times10^{-16}$ for the PGS predictor. Adjusted $R^2$ is the corrected goodness of model fit. Values in square brackets represent 95% confidence intervals. Comparison $R^2$ values are those metrics from the original studies generating the PGS panels. Model covariates or predictors used in the comparison metrics are mentioned in brackets next to the values.
